# Constructing a Literature-Derived Database for Benchmarking Polygenic Risk Score Construction Methods with Spectral Ranking Inferences

**DOI:** 10.64898/2026.03.01.26347258

**Authors:** Chris Sebastian, Mengxin Yu, Jin Jin

**Affiliations:** Department of Mathematics, University of Pennsylvania, Pennsylvania, PA 19103; Department of Statistics and Data Science, Washington University in St. Louis, St. Louis, MO 63130; Department of Biostatistics, Epidemiology and Informatics, Perelman School of Medicine, University of Pennsylvania, Pennsylvania, PA 19103

**Keywords:** Polygenic Risk Score (PRS), Spectral ranking inference, Meta-aggregation of literature data, Method benchmarking database, Uncertainty quantification

## Abstract

Polygenic risk scores (PRSs) have emerged as a valuable tool for genetic risk prediction and stratification in human diseases. Over the past decade, extensive methodological efforts have focused on improving the predictive power of PRS, leading to the development of numerous methods for PRS construction. Benchmarking these various methods thus becomes an essential task that is crucial for guiding future PRS applications. While studies have benchmarked subsets of these methods on specific phenotypes and cohorts, the resulting evidence remains fragmented, with a lack of work that comprehensively assess the relative performance of the various PRS methods. In this study, we addressed this gap by systematically constructing a PRS method benchmarking database synthesizing published results from 2009 to 2025. We applied a spectral ranking inference framework with uncertainty quantification to rank 14 PRS methods that had been adequately compared against each other in the literature. We constructed rankings using two complementary sources: original method-development studies and applications/benchmarking studies. While the highest-ranked methods (LDpred2 and AnnoPred) and the lowest-ranked method (C+T) were consistently identified from both sources, the relative ordering of most methods showed moderate variability. We further constructed phenotype-specific rankings, providing more detailed insights into the robustness and phenotype-specific strengths of individual methods. Collectively, the overall and phenotype-specific rankings of the PRS methods, along with the curated benchmarking data from the literature, provide a dynamic and practical reference database that can continuingly be updated with emerging new PRS methods and published benchmarking results to guide future PRS applications.

## Introduction

Polygenic risk scores (PRSs), derived from genome-wide association studies (GWAS), are increasingly recognized as quantitative measures of genetic liability with great potential for facilitating disease risk stratification, personalized risk management, and clinical decision-making[1–3]. Recent studies have highlighted the clinical utility of PRS when integrated with complementary data modalities, such as electronic health records (EHR)[4–7] and multi-omic profiles[8–10]. A wide range of methods have been proposed for PRS construction, with many approaches focusing on the use of GWAS summary-level association statistics, which are far more widely available than individual-level GWAS data. Early approaches, such as LD Clumping and p-value Thresholding (C+T)[11,12] —also known as P+T —and related implementations such as PRSice[13], were widely adopted due to their computational efficiency, algorithmic simplicity, and ease of interpretation. More advanced methods were subsequently developed, including Bayesian approaches (e.g., LDpred2[14], PRS-CS[15], and SBayesR[16]) and penalized regression-based methods (e.g. lassosum2[17]). By explicitly accounting for complex correlation structures among millions of genetic variants, these methods have demonstrated improved predictive performance over earlier, model-free approaches.

To date, over 30 methods have been proposed for PRS model development, with individual method-development papers often claiming superiority over existing approaches for specific phenotypes or data scenarios. The relative performance of PRS methods is known to depend on multiple factors, including how well the distribution assumption approximates the true effect-size distribution of genetic variants of a phenotype, algorithm robustness, and other factors[3]. In addition to the original method papers, which typically benchmark newly proposed methods against a subset of existing methods on a limited set of phenotypes for illustrative purpose, subsequent application and benchmarking studies have evaluated different subsets of PRS methods across a broader range of phenotypes and datasets. Together, results reported in both method-development and applied benchmarking papers serve as valuable but fragmented resources for understanding PRS method performance and informing method selection in future applications. However, there remains a lack of synthesis efforts that comprehensively summarize the relative performance of PRS methods across diverse phenotypic categories and data settings. Integrating findings from the existing literature using appropriate analytical frameworks could provide a more systematic assessment of method robustness and phenotype-specific performance patterns. Moreover, a stratified investigation based on the type of paper: either the original method papers or the independent applied and benchmarking studies, may further offer a balanced evaluation and provide additional insights.

In this study, we constructed PRS method rankings by synthesizing all benchmarking results reported in the published literature using a spectral ranking inference approach with uncertainty quantification. We identified 14 methods for GWAS summary data-based PRS construction that had been sufficiently compared against each other on complex human phenotypes. Method comparison results were collected from 35 PubMed publications published between 2009 and 2025. These literature-derived data posed a series of challenges for synthesis analysis, including sparsity of pairwise method comparisons and incomparability of results across applications due to differences in evaluation metrics, target phenotypes, and study cohorts. To address these challenges, we adopted a recently proposed spectral ranking inference approach that enables statistically principled aggregation of sparse and heterogeneous comparison data while providing confidence intervals (CIs) for inferred ranks of the PRS methods. Our analysis identified several methods that were consistently ranked at the top (LDpred2 and AnnoPred) or bottom (LDpred2-inf and C+T) with high confidence, based on evidence drawn either from original method-development papers or from subsequent application and benchmarking papers. Meanwhile, the remaining methods had rankings with largely overlapping CIs, suggesting similar overall performance. From the method paper-based rankings, we observed two clusters of methods where one (including LDpred2, AnnoPred, LDpred2-auto, LDpred-funct, and DBSLMM) outperformed the other (including PRS-CS, SCT, LDpred, PRS-CS-auto, LDpred2-inf, and C+T). We also observed a notable pattern that the rankings tended to be higher for methods proposed more recently. On the other hand, with a larger number of applications across a wider range of phenotypes, the rankings based on application papers gave much narrower CIs for most methods and the methods’ ranks were less related to their publication dates. While most methods’ ranks were comparable between rankings derived from method-development studies and those based on applied or benchmarking studies, a small number of methods (including SCT and DBSLMM) showed notable differences in rank between the two rankings. We further conducted phenotype-specific summary analyses, generating rankings stratified by phenotype. These analyses revealed substantial variability in methods’ relative performance across phenotypes and provided more detailed insights into the robustness and phenotype-specific strengths of individual PRS methods. Together, the overall and phenotype-specific ranking results, along with the curated literature-based benchmarking data, provide a comprehensive reference database for benchmarking PRS methods and guiding method selection in future PRS development and applications across diverse complex human phenotypes.

## Methods

### Problem Setup and Data Preparation

We conducted a comprehensive PRS method performance ranking with uncertainty quantification based on benchmarking results collected from the literature. Specifically, we first identified 14 methods for GWAS summary data-based PRS construction which were sufficiently compared against others. These methods include: C+T[12,18], LDpred[11], lassosum[19], AnnoPred[20], PRS-CS-auto[15], PRS-CS[15], SBayesR[16], SCT[21], DBSLMM[22], LDpred2-inf[14], LDpred2-auto[14], LDpred2[14], LDpred2-funct[23], lassosum2[17], sorted by publication date (from earliest to latest). These are commonly implemented methods in the standard PRS construction setting where a single GWAS summary dataset is available for model training, referred to as the single ancestry setting. Other categories of PRS methods, such as those in the multi ancestry setting where ancestry-stratified GWAS summary data is available from more than one ancestry population for model training, those in the admixed population setting aiming to improve PRS performance on admixed populations, and cross-trait methods aiming to improve PRS performance by borrowing information across related traits, have mostly not been sufficiently compared in the literature and are thus not included in our analysis. Finally, we collected 28 papers from PubMed published between 2009 and 2025 that reported method comparison results among the 14 methods based on 536 applications across 108 distinct phenotypes and 5 superpopulations (Figure 1). These phenotypes span across several categories of human complex traits including binary clinical endpoints and disease outcomes, continuous physiological traits (e.g., blood lipids, hematological traits), and behavioral traits.

**Figure 1:**
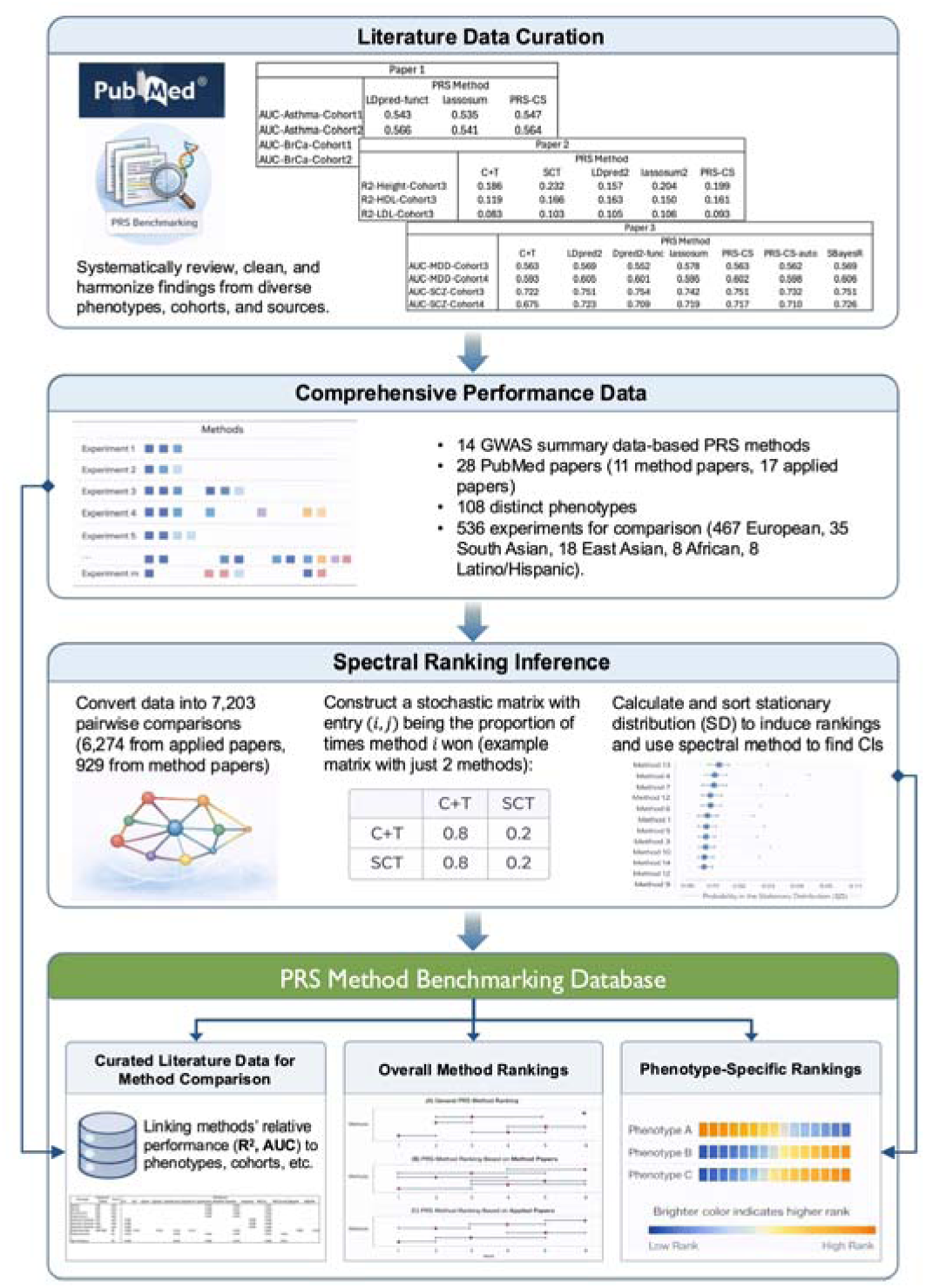
An overview of the study pipeline. The pipeline started from performing a systematic review of the relevant literature, extracting and curating the relative performance data, to generating model ranks using spectral ranking inference. The final product is a PRS method benchmarking database consisting of three components: (1) curated literature data (R^2^ and AUC values) linking methods’ relative performance to specific phenotypes and cohorts; (2) overall method rankings based on all papers, the original method-development papers only, and the applied and benchmarking papers only; and (3) phenotype-specific rankings of the PRS methods.

The data collected came from two categories of paper sources, the first being the original method-development papers which introduced the PRS methods, and the second being the applied and benchmarking papers that studied the relative performance of multiple PRS methods on specific phenotypes or cohorts. Each method paper, besides the one that introduced the first method, C+T[12], had a data analysis section comparing the proposed method with the previously proposed methods or a subset of them on a set of phenotypes for illustrative purpose. These comparisons were typically performed based on *R*^2^ performance on predicting continuous phenotype values or Area under the ROC Curve (AUC) performance on classifying binary outcomes. We centralized these reported findings into a data table, with the rows corresponding to the various experiments on the same or different phenotypes and the columns corresponding to the PRS methods. The data collection was largely similar for the applied and benchmarking papers. Most papers were benchmarking efforts which had the comparison results structured in a similar way to the method papers: testing a series of methods against a series of phenotypes. Meanwhile, some other papers were testing the methods’ performance on just one phenotype. In these papers, the comparisons were done not by phenotype but by some other differentiating factors, such as study cohort. Thus, in these datasets, the columns still correspond to the methods being tested, but the rows now correspond to experiments on the different cohorts. In both types of papers, there were often real data applications accompanied by simulated studies. We include results only from real data applications but not simulation studies because first, we aim to rank the methods based on real world performance, and second, simulation studies, which often generate synthetic data based on the assumed model assumptions, tend to favor the proposed methods. The collected data has a unique structure in that method comparisons are often conducted among subsets of methods on different phenotypes and cohorts, making the data table relatively sparse with missing entries and non-comparable rows. These unique data features cannot be easily handled by simple method benchmarking approaches such as taking an average across all applications. On the other hand, a statistically principled approach that can robustly integrate results across diverse studies can provide a better solution to our task.

### Spectral Ranking Inference

The spectral inference method, described in Fan et al. 2025[24], allows for the estimation of ranks with CIs for a set of objects that are being compared. The method begins by having a set of objects, numbered 1 through *n*, which we will call *Ω* . Additionally, there must exist a collection of subsets of *Ω*, which we will call comparison sets. There must also exist a criterion for choosing the most favorable object from any given subset, which we will call the “winner” of the subset. Having this collection of subsets and their respective winners allows you to construct an *n* × *n* stochastic matrix representing the objects in *Ω*. Specifically, for non-diagonal entries, entry *(i, J)* is an increasing function of the proportion of times the *i* -th object in *Ω* is a winner of a comparison set in which the *J*-th object is also a member. Each diagonal entry is defined as one minus the sum of the other (appropriately scaled) entries in its row, ensuring that every row sums to 1 and contains no negative elements. With the stochastic matrix constructed, one can then calculate the stationary distribution of the matrix by finding the top-left eigenvector of the stochastic matrix. The estimated ranking of the objects is now defined by sorting the objects in descending order of their probability in the stationary distribution. The intuition being that those which have the largest probabilities are the ones winning the most comparison sets and thus should be ranked highest[25,26]. Through further calculations with the spectral method, we can derive uncertainty quantifications in the form of 95% CIs of the estimated rankings using the bootstrap method discussed in Fan et al. 2025[24].

The spectral inference method works well in our problem setting as it aggregates performance across the multitude of comparisons found spanning the literature. Furthermore, the production of CIs allows for a richer analysis and comparison of methods within and between different rankings. In the context of our study, the set *Ω* was the 14 PRS methods we chose to study. When constructing the comparison sets, we defined entry *(i, J)* to be an increasing function of the proportion of times the *i*-th object in *Ω* was a winner of a comparison set in which the *J* -th object was also a member. This means the stochastic matrix did not consider the relative performance of two methods in a comparison set if neither was a winner of the comparison set. Hoping to avoid such situations and maximize the information obtained from the given data, we made all our comparison sets of size two, i.e., conducting inference based on pairwise comparison of the method performance. To complete this data reconstruction, we decomposed each comparison, for example, method comparison on a specific phenotype/cohort, which compared *k* ≥ 2 methods, into 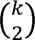 pairwise comparison sets. For each comparison set, we defined the winning method as the method which had a higher prediction *R*^2^ or AUC value. If two methods had the exact same metric score, then no winner was declared, and that data point was not included in the calculations (Figure 1). This was not a common occurrence, though. As an example, consider an imaginary dataset containing the AUC performance of LDpred2, lassosum2, and PRS-CS for developing a PRS model for classifying the binary Type 2 Diabetes (T2D) status. Suppose there are 3 pairwise comparisons, with example AUC values in parathesis: [LDpred2 (.8) vs lassosum2 (.7)], [LDpred2 (.8) vs PRS-CS (.65)], and [lassosum2 (.7) vs PRS-CS (.65)]. The winner of each comparison, respectively, would be LDpred2, LDpred2, and lassosum2.

## Results

### The General Ranking of Methods

We constructed and compared three sets of rankings based on method performance reported in the literature. Detailed information on the extracted literature data is summarized in Supplementary Table 1. We first constructed the overall rankings by combing data from both the original method papers and the applied/benchmarking papers. Figure 2(A) presents the overall ranking results: the methods were listed chronologically based on the publication date of their method papers, with the very first (C+T) being at the top of the method axis and the newest (lassosum2) being at the bottom. LDpred2 and AnnoPred are ranked the highest with tight CIs that showed significant superiority over most of the other methods except lassosum2, SCT, and LDpred-funct. C+T, LDpred, and LDpred2-inf are ranked the lowest with similarly high degrees of confidence that clearly distinguish them from higher ranked methods. The other 9 methods tend to cluster in the middle of the ranking and have wider CIs, suggesting a similar performance overall. A large number of comparisons conducted for a method did not necessarily lead to a narrow CI for the method’s rank. For example, SBayesR had the third largest number of comparisons but also had a rank with a second widest CI (Figure 2(A)). The occurrence of the wide, overlapping CIs of rankings across most methods may be due to the large variation in genetic architectures across different phenotypes (e.g., heritability, effect size distribution of genetic variants) and data settings (e.g., GWAS sample size, ancestral population) that could lead to highly heterogenous relative performance between different methods.

**Figure 2:**
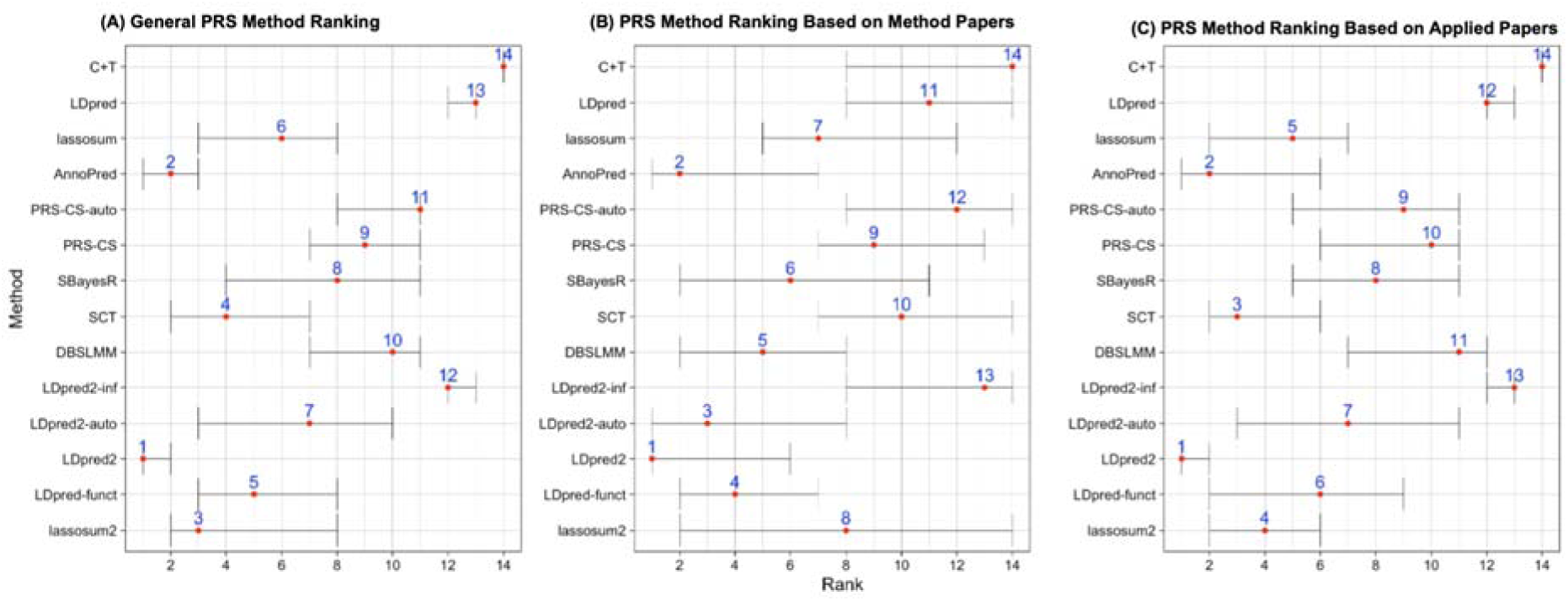
Spectral method inferred rankings of 14 PRS methods based on benchmarking results reported in the literature. **(A)** General method rankings estimated using data from all papers. **(B)** Method rankings estimated based on data from the original method-development papers. **(C)** Method rankings estimated based on data from the subsequent application and benchmarking papers. The red dots and grey bars show point estimates and 95% CIs, respectively, of the rankings. The methods are ordered chronologically (by publication date of their method papers) with C+T being the first method and lassosum2 being the latest.

We further generated two separate rankings based on results reported in the original method papers and in the applied/benchmarking papers, respectively. Detailed method comparison results, including the number of wins and losses of each method against others, are reported in Supplementary Table 2. Figure 2(B) shows the ranking produced based only on results collected from the method papers. Given that the experiments conducted in the method papers were typically for illustrative purpose, comparisons were conducted only on a limited number of phenotypes and cohorts, which could partially explain the wider CIs of the rankings compared to those in Figure 2(A). Even so, we can observe approximately two clusters of methods where one (including LDpred2, AnnoPred, LDpred2-auto, LDpred-funct, and DBSLMM) outperforms the other (including PRS-CS, SCT, LDpred, PRS-CS-auto, LDpred2-inf, and C+T). Three other methods, SBayesR, lassosum, and lassosum2, show rankings with wider CIs that overlap with both clusters. A notable pattern is that the rankings tend to be higher for methods proposed more recently: most of the lower-ranking methods are those proposed earlier (top half of the method axis, C+T through SBayesR), while most of the higher-ranking methods are generally those proposed later (bottom half, SCT through lassosum2). Such a general trend is consistent with the claim in the method papers that newer methods perform better, with a few exceptions: AnnoPred ranking second, LDpred2-inf ranking thirteenth, and the newest method lassosum2 ranking 8^th^, but with wide CIs that overlap with that of many other methods.

As a comparison, we further conducted method ranking only using data from application and benchmarking papers (Figure 2(C)). This serves as a control group with objective method benchmarking on a wider range of phenotypes. Detailed method comparison results, including the number of wins and losses of each method against others, are reported in Supplementary Table 3. Given that these applied papers report a much larger number of comparisons (6,274 compared to 929 from method papers), the CIs are generally tighter, allowing for more statistically significant differentiations to be made. This leads to some similar (as compared to rankings based on method papers in Figure 2(B)) but more significant rankings. For example, LDpred2-inf, LDpred, and C+T, which are ranked among the worst methods based on method papers, are now still ranked among the worst methods, but with higher certainty. LDpred2 and AnnoPred remain the top two methods with higher certainty. Among the remaining methods in the middle, some have similar rankings as before but with narrower CIs, such as SBayesR (6 to 8), lassosum (7 to 5), PRS-CS (9 to 10), and PRS-CS-auto (12 to 9). However, three methods have a significant change in ranking positions: SCT goes up from 10^th^ to 3^rd^ and DBSLMM goes down from 5^th^ to 11^th^ (both having narrower CIs), while lassosum2 goes up from 8^th^ with a wide CI to 4^th^ with a much narrower CI. These wide swings in performance are likely due, in part, to the greater variability in phenotypes present in the applied paper dataset than in the method paper dataset. The method papers typically tested the same few phenotypes (e.g. cancers and blood lipid traits) and might not provide a holistic evaluation of a method’s general performance. From the applied paper-based ranking, some of the earliest proposed methods, such as lassosum, AnnoPred, and SCT, are ranked in the top half of methods with high confidence, while DBSLMM, a more recently proposed method, is ranked in the lower half of methods. These results suggest that, with a larger number of applications across a wider variety of phenotypes, the overall performance of PRS methods seem to be less related to their publication date (compared to the findings from method paper-based ranking in Figure 3(B)). Despite some apparent differences, the two rankings in Figure 2(B) and Figure 2(C) do assign the same methods to be ranked first and second, as well as the same methods ranked last and second to last. Note that, while LDpred2-inf and LDpred2-auto were listed among the recently proposed methods, they were proposed along with LDpred2 as a simple version and a tuning-parameter-free version, respectively, of LDpred2. LDpred2-inf was expected to have suboptimal performance, which may explain its low rank in both rankings.

**Figure 3:**
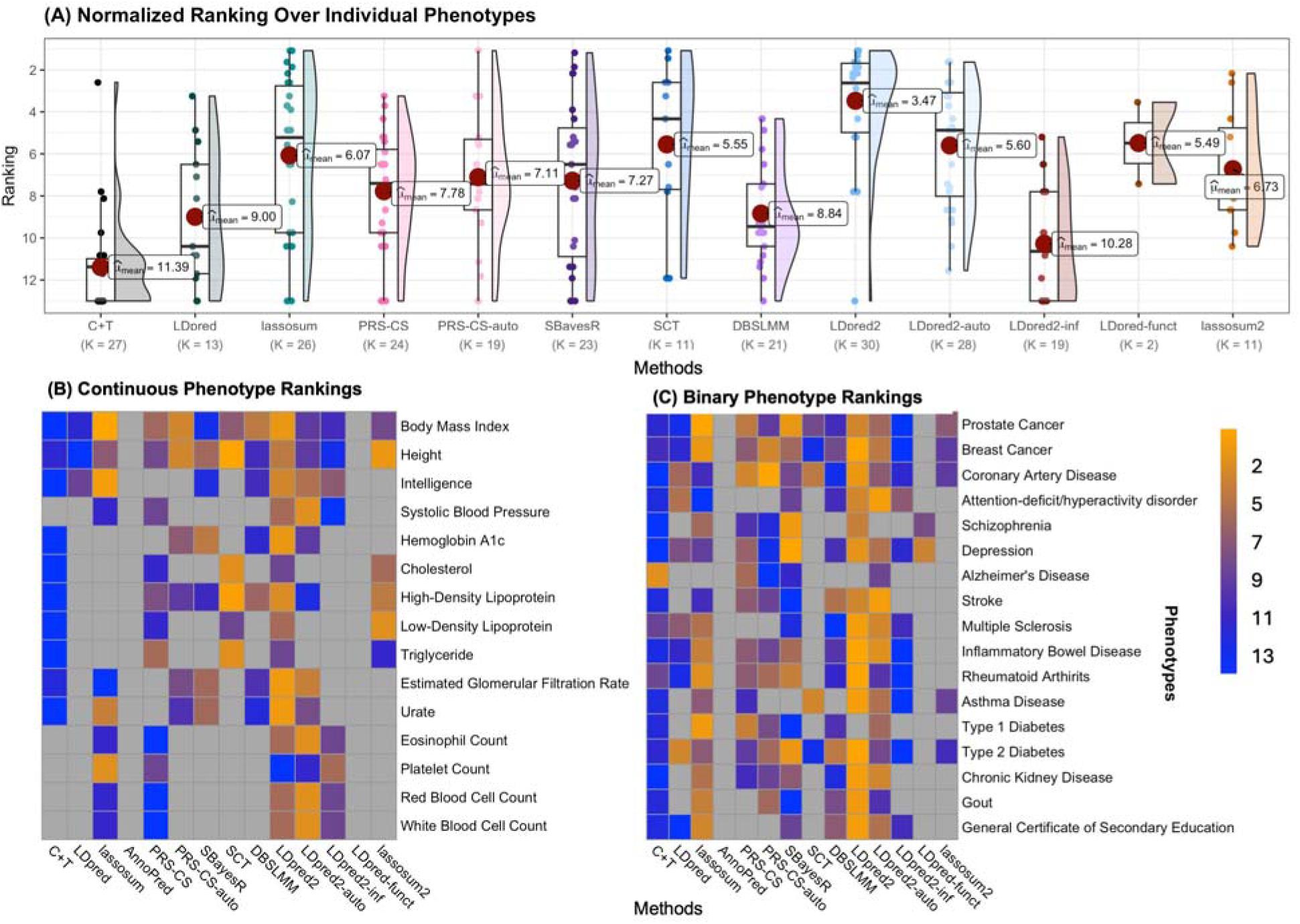
Phenotype-specific rankings of PRS methods. **(A)** Half violin plot showing normalized ranking across eligible phenotypes. The dots in dark red show the average inferred rankings of each method across all eligible phenotypes. The expression (K = x) under each method shows the number of phenotypes the method passed the filtering check and was ranked for. **(B)** A heatmap showing the ranks the methods achieved when tested against given continuous phenotypes. **(C)** A heatmap showing the ranks the methods achieved when tested against given binary phenotypes. The color ranges from orange, representing high ranks, to blue, representing low ranks. Grey cells occur when the method was not ranked on the specific phenotype because it was removed during the described filtering process. AnnoPred failed to be compared at least 10 times for any phenotype and was thus omitted from the phenotype-specific comparison. Details of the normalized ranks for continuous and binary phenotypes are summarized in Supplementary Tables 5 and 6, respectively.

Overall, the two sets of rankings based on separate data sources from method papers and applied papers provide complementary observations of PRS method performance. The highest and lowest ranked methods are relatively consistent between the two rankings but with the applied rankings providing narrower CIs: possibly due to the much larger number of experiments included. For the majority of the methods, the CIs of their rankings are wide, showing no significant difference across most of the method pairs. Nonetheless, there does exist statistically significant difference in overall method performance, especially within the rankings based on results reported in applied papers. These rankings provide researchers with unique perspectives about PRS method performance with diverse benchmarking summary of the PRS method performance based on different literature sources, which could help better inform the choice of PRS methods for future PRS construction.

### Phenotype-Specific Analysis

The rankings reported in Figure 2 were calculated by aggregating benchmarking results from the literature across a total of 108 distinct phenotypes. These rankings provide an overall assessment of the relative performance of PRS methods: reflecting their robustness and general applicability across diverse genetic architectures and application settings. However, the relative performance of PRS methods is known to be phenotype dependent. Thus, we further investigate phenotype-specific rankings as a complementary analysis, which could provide insights into method performance within individual trait categories and inform method selection for future PRS model development targeting on specific phenotypes.

To construct phenotype-specific rankings, a naive approach is to partition the dataset into phenotype-specific sub-datasets and calculate ranking on each sub-dataset. This approach, however, presents a unique challenge, as some methods had very few comparisons and thus their rankings were uninformative or simply misleading. This is because there were many phenotypes for which one method had less than 5 comparisons but always won. This situation led to a stationary distribution that put all its mass on the method which had a 100%-win rate and 0% on all other methods. This created two problems: (1) the other 13 methods could not be ranked, and (2) it is unconvincing that this method ranked first when it was only compared 5 times. To address this issue, we filtered out any method which had fewer than 10 comparisons for a given phenotype. This ensured that all the methods present after filtering had enough comparisons for a given phenotype to produce a meaningful ranking. During the process, AnnoPred failed to be compared at least 10 times for any phenotype and was thus omitted from the phenotype-specific comparison. This approach largely solved the stated issue. However, there were still a few instances where a method won all its comparisons. Given that the method had over 10 comparisons, though, there was enough data to believe this wasn’t an anomaly. However, the 0% win rate assigned to other methods would make the ranking impossible for all other methods. To ensure all methods could be ranked, we thus removed this top method from the sub-dataset, labeled it as the top ranked method, and then re-ran the ranking calculation on the remaining methods. This had to be an iterative process, because once you removed the highest ranked method from the dataset, the second highest ranked method could have been undefeated amongst all remaining methods. Thus, we iterated this process until a final ranking across all methods was achieved.

After method filtering, each phenotype *p* had a ranking of a subset of the 13 methods (AnnoPred was filtered out due to insufficient comparisons), which we will refer to as *M_p_*. The cardinality of *M_p_* varies from 0 to 13 depending on *p* because each method had a variable number of comparisons for any given phenotype *p*. The varying cardinality makes it difficult to compare rankings across phenotypes because a ranking of first out of two methods should hold less weight than a ranking of first out of 13 methods. To address this issue, we performed a normalization on ranking across methods for each phenotype. Specifically, if a method achieved a rank *r*, we defined its final ranking *r_f_* as

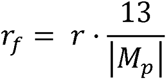

This normalization factor 13/|*M_p_*| penalizes rankings which had a small number of methods present. The more methods present, the smaller the normalization factor is. At its smallest, the normalization factor is 1 and has no impact on the ranking, allowing the best method to achieve a final ranking of 1. At its largest, the normalization factor is 6.5 (when *M_p_* contains the smallest possible number of methods, 2), making the best method’s final ranking 6.5. This ensures that methods are only listed as ranking highly if there is an adequate amount of data to support that conclusion. Information on the number of comparisons by phenotype is summarized in Supplementary Table 4. The final rankings after normalization across 15 continuous and 17 binary phenotypes for which PRS methods were adequately compared are summarized in Figure 3.

We observe that the average method performance across phenotype in Figure 3(A) yields a very similar ranking to the overall ranking in Figure 2(A). As expected, the large variation in most methods’ ranks across phenotypes suggests that the generally “worst” method could outperform the generally “best” method when looking at a specific phenotype. For example, C+T was ranked with high certainty as the worst method across the applied papers, but when looking at Figure 3(C) one can find it is ranked 2^nd^ for Alzheimer’s Disease. While LDPred2, which was ranked with high certainty as a top 2 method, was ranked 13^th^ for Platelet Count (Figure 3(B)). These phenotype-specific rankings presented in Figures 2(B) and 2(C) clearly articulate the robustness and specialty (on phenotypes highlighted with light orange) of each PRS method. For example, we can observe a consistent superiority and high robustness of LDpred2 across most phenotypes considered, while SBayesR appeared to be a suitable choice for training PRS models for Schizophrenia and Depression, both being highly polygenic phenotypes that have widespread causal genetic variants across the genome. In general, these phenotype-specific rankings, alongside the overall rankings, synthesizing benchmarking efforts from the literature, provide a comprehensive landscape of PRS method comparison that could inform future PRS training. Researchers can refer to the relevant part of the phenotype-specific rankings, along with the raw benchmarking data we extracted from the literature reporting detailed method comparison results by phenotype, population, and cohort (Supplementary Table 1), for choosing PRS methods that could be most promising for specific phenotypes of interest.

## Discussion

Understanding the relative performance of computational methods through systematic synthesis of the existing literature can yield important insights into their real-world utility and inform future methodological development and implementation. In the context of polygenic risk score (PRS) modeling, new methods continue to emerge, while comparative evaluations are typically conducted on limited subsets of methods across disparate studies. As a result, the available benchmarking evidence remains fragmented. Establishing a comprehensive benchmarking resource that integrates all available evidence from the literature is therefore essential for thoroughly investigating the relative performance of PRS methods and guiding their future applications. Our study leads such an effort by systematically establishing a PRS method benchmarking database, summarizing the relative performance of PRS construction methods with available benchmarking data from the literature. We curated publicly available benchmarking results for 14 GWAS summary data-based PRS construction methods from 28 PubMed publications and applied a statistically rigorous spectral ranking inference approach with uncertainty quantification to integrate these sparse and heterogeneous comparison results. We derived PRS methods’ rankings for their overall performance based on (1) all available publications, (2) original method-development papers only, and (3) applied and benchmarking papers only. Although the current data size (6,274 pairwise comparisons in total, with an average of 448 comparisons per method) yielded relatively wide CIs for the ranks of many methods, several methods consistently clustered near the top (LDpred2 and AnnoPred) or the bottom (C+T and LDpred2-inf) across different rankings. Finally, we ranked methods’ performance by individual phenotypes, revealing more detailed and highly heterogeneous rankings across different categories of phenotypes. No clear trend of method ranks by publication date was observed, suggesting that the newer methods do not necessarily outperform earlier ones. However, the large variability in ranks and failure to differentiate ranks among most of the methods suggest that there are no “best” methods, and that future method development could continue being specialized for specific genetic architectures or introducing more flexibility in handling the genetic nuances across phenotypes.

Given current data availability, our PRS method ranking is restricted to methods that construct PRS using a single GWAS summary-level dataset, without incorporating population heterogeneity or conducting multi-trait training. There are other recently proposed methods in this category but were not considered due to insufficient benchmarking data to support reliable ranking. As more benchmarking data become available, these methods can be integrated into our existing ranking database for single-ancestry PRS methods[27–29]. Beyond this setting, a growing body of work has focused on extending standard PRS modeling frameworks to improve predictive performance and generalizability, such as multi-ancestry methods[30–33], multi-trait methods[34], and methods incorporating admixed or local ancestry information[35–37]. As more benchmarking results for these recent advanced methods continue to emerge, our ranking framework and database can be naturally expanded to establish rankings for these additional categories of PRS methodologies[38].

The PRS method rankings presented in this study should be interpreted with caution. First, while our analyses have synthesized benchmarking results from the literature, the phenotypes covered are still limited in number, suggesting that only a partial insight is available from our current ranking database. Nevertheless, we anticipate that a more comprehensive analysis will be feasible in the near future, given recent large-scale benchmarking efforts reported in preprints that evaluated relative performance of PRS methods across a broader range of phenotypes[39,40]. Second, our ranking framework provides a robust way to summarize results across different studies and phenotypes, for which the predictive performance metrics such as R^2^ or AUC are not directly comparable, and the comparison tables are often sparse. While the spectral inference approach allows a robust ranking under these conditions, it characterizes relative ordering in rather than the magnitude of performance differences in specific metrics. Consequently, such rankings, especially the phenotype-specific rankings, should be interpreted in conjunction with the corresponding R^2^ or AUC values in our collected literature database to more fully inform future method implementation. Third, all comparisons in our current analysis were weighted equally across studies. However, different studies may contribute differently (e.g., due to varying sample sizes) with varying uncertainty in reported R^2^ or AUC values. We currently do not take into account the uncertainty in the reported prediction R^2^ or AUC by assigning study-specific weights, mainly because such information is unavailable in most studies. With the availability of future benchmarking studies that report such uncertainty measures, our ranking pipeline can be extended to incorporate study-specific weights, such as weights proportional to the inverse of variance of the estimated R^2^ or AUC values, to further refine the rank inference. In addition, our current rankings focus exclusively on predictive accuracy and do not consider computational efficiency or computational resource requirements, which are also important practical considerations for PRS method selection. Finally, our analyses primarily focused on complex human phenotypes, while other phenotypes classes, such as molecular traits and imaging traits, that also benefit from PRS modeling[41–43], could be examined as well[41].

In summary, our PRS method benchmarking database, including the curated literature data on methods’ relative performance (Supplementary Table 1) and generated method rankings (Figures 2 and 3), provide a valuable reference resource for PRS method comparison and implementation across a broad range of human traits and diseases. We further identify a series of directions for extending this resource, highlighting the need for future efforts to continuingly update the constructed database as new methods and benchmarking studies emerge. Such efforts will enable the development of a dynamic and practical reference framework to guide PRS method selection and implementation in future research and applications. More broadly, we hope that the PRS application of our general analysis framework for curating and synthesizing literature evidence to construct benchmarking databases for computational methods could provide insights into the feasibility of extending the framework to other domains where comprehensive benchmarking of computational methods is needed.

## Data Availability

Data can be found in the supplementary data and the github which contains code and raw data.

https://github.com/codeitchris/PRS-Ranking-Paper-Code-and-Data/tree/main

## Competing interests

The authors declare no conflict of interest.

## Data and code availability

All code used for data cleaning, graph making, ranking calculation, and the collected datasets on method performance are available at https://github.com/codeitchris/PRS-Ranking-Paper-Code-and-Data.

## Study Funding

This work was supported by the National Institutes of Health [R00 HG012223 to J.J., R35GM157133 to J.J.].

## Author Contributions

C.S. and J.J. conceived and designed the project. C.S. conducted data analyses under the supervision of J.J. C.S. and J.J. drafted the manuscript, and M.Y. provided comments. All co-authors reviewed and approved the final version of the manuscript.

## Notes

### Competing Interest Statement

The authors have declared no competing interest.

### Funding Statement

This work was supported by the National Institutes of Health [R00 HG012223 to Jin Jin, R35GM157133 to Jin Jin].

